# Blood group A Secretors are associated with a higher risk of COVID-19 cardiovascular disease complications

**DOI:** 10.1101/2020.12.19.20248172

**Authors:** TJ Mankelow, BK Singleton, PL Moura, CJ Stevens-Hernandez, NM Cogan, G Gyorffy, S Kupzig, L Nichols, C Asby, J Pooley, G Ruffino, F Hosseini, F Moghaddas, M Attwood, A Noel, A Cooper, D Arnold, F Hamilton, C Hyams, A Finn, AM Toye, DJ Anstee

**Author notes:** Correspondence: Prof. David Anstee, BITS, NHSBT, North Bristol Park, Filton, Bristol BS34 7QH, United Kingdom;, Dr Ashley Toye, School of Biochemistry, University of Bristol, Bristol BS8 1TD, United Kingdom.

## Abstract

The SARS-CoV-2 virus causes COVID-19, an infection capable of causing severe disease and death but which may also be asymptomatic or oligosymptomatic in many individuals. While several risk factors, including age, have been described, the mechanisms of this variation are poorly understood. Several studies have described associations between blood group and COVID-19 severity, while others do not. Expression of ABO glycans on secreted proteins and non-erythroid cells is controlled by a fucosyltransferase (FUT2). Inactivating mutations result in a non-secretor phenotype which is known to protect against some viral infections. We investigated whether ABO or secretor status was associated with COVID-19 severity. Data combined from healthcare records and laboratory tests (n=275) of SARS-CoV-2 PCR positive patients hospitalised with COVID-19, confirmed higher than expected numbers of blood group A individuals compared to O (RR=1.24, CI 95% [1.05,1.47], P=0.0111). There was also a significant association between group A and COVID-19-related cardiovascular complications (RR=2.56, CI 95% [1.43,4.55], P=0.0011) which is independent of gender. Molecular analysis of phenotype revealed that group A patients who are non-secretors are significantly less likely to be hospitalised than secretors. In a larger cohort of 1000 convalescent plasma donors, among whom the majority displayed COVID-19 symptoms and only a small minority required hospitalisation, group A non-secretors were slightly over-represented. Our findings indicate that group A non-secretors are not resistant to infection by SARS-CoV-2, but they are likely to experience a less severe form of its associated disease.

**Key Points:**

1. **Blood group type A is associated with an increased risk of cardiovascular complications in COVID-19 patients.**
2. **FUT2 “non-secretor” status reduces the risk of severe COVID-19 outcomes in patients with blood group A.**

## Introduction

Genetic diversity among members of animal species including *Homo sapiens* is essential for their survival in response to newly emergent and evolving pathogens.^1^ Human blood group antigens are among the first polymorphic structures encountered by viruses and bacteria upon airborne contact with respiratory, gastrointestinal and urinogenital mucosal surfaces.^2^ The carbohydrate antigens of the ABO and Lewis blood group systems are found on mucosal surfaces where their presence is controlled by a fucosyltransferase (FUT2).^3,4^ In the presence of active *FUT2* A, B, H and Le^b^ antigens can be expressed on mucosal surfaces. Individuals with this phenotype are known as secretors^5,6^. In individuals lacking active *FUT2*, known as non-secretors, only the Le^a^ antigen can be expressed^5^. There are several well studied interactions between host cells and both bacteria (*Helicobacter pylori, Vibrio cholera*) and viruses (noroviruses, rotaviruses) which are known to depend on the presence of these antigens ^7,8^. In particular, it is clearly established that common strains of norovirus and rotavirus fail to infect non-secretors.^9-12^

A novel coronavirus [SARS-CoV-2 (severe acute respiratory syndrome coronavirus 2)] emerged in China in December 2019, causing a pandemic of severe respiratory disease, known as coronavirus disease (COVID-19)^13^. Currently, more than 71 million people have been infected worldwide and, of those, more than 1.6 million have died as a result of the disease. It was apparent from the earliest studies that disease severity in individual patients varied considerably, ranging from asymptomatic infection to fatal illness.^14^ Zhao et al. provided the first evidence of an association between blood group polymorphisms and with disease severity.^15^ In a study of over 2,000 infected patients from Wuhan, China, it was noted that group A phenotype was found more frequently than expected in patients with COVID-19, whereas group O occurred less commonly than expected in the general population, suggesting that group A individuals are at greater risk from COVID-19 than those of group O. This observation led us to undertake a study of the association of blood group polymorphisms (ABO and secretor status) with COVID-19 severity in hospitalised patients in Bristol, UK.

Here we report a significant increased frequency of hospitalisation for blood group A compared to blood group O patients with COVID-19. This was accompanied by a significantly higher rate of respiratory failure at admission but had no significant association with length of stay or patient death rate. Importantly, we observe a trend between blood group A and cardiovascular complications, which we confirm in a separate cohort of hospitalised COVID-19 patients. Investigation of the role of secretor status in relation to disease severity revealed that COVID-19 incidence in group A non-secretors was much lower than would be expected if the absence of active FUT2 had no impact on disease, within the hospitalised patient context. Moreover, this effect was specific to blood group A and was not observed in patients with blood group O

## Methods

### Epidemiological Surveillance

A retrospective cohort analysis of COVID-19 infected individuals was undertaken as part of an audit on adult patients hospitalised at North Bristol NHS Trust with COVID-19 infection, was approved by the North Bristol NHS Trust Audit and Research Ethics Committee.. Adults admitted from 27 March to 27 July 2020 were identified by searching the Laboratory Information Management System (LIMS) database (Clinisys, WinPath Enterprise). The inclusion criteria were a positive PCR result for SARS-CoV-2, using the established Public Health England reverse transcriptase polymerase chain reaction (RT-PCR) assay in use at the time and the requirement for hospitalisation. Clinical records were then reviewed to determine patient demographics, pre-existing co-morbidities and blood group. Outcomes were assessed 30 days following admission, including the requirement for cardiovascular and respiratory support. A random number generator was used to select 10% of records for review to ensure data accuracy.

### Blood samples

Access to, and research on healthy donor and COVID-19 patient samples was undertaken using Health Research Authority (HRA) ethical approval, which was reviewed and approved by Leeds West Research Ethics Committee (REC No. 20/YH/0168). This includes accessing blood samples and clinical outcomes collected by the DIagnotic and Severity markers of COVID-19 to Enable Rapid triage (DISCOVER) study of hospitalised COVID-19 patients and outcomes between 1^st^ April and 1^st^ October 2020. The DISCOVER Study is an observational cohort study of patients with either a PCR positive test or COVID-19 symptoms at North Bristol NHS Trust. DISCOVER samples were collected under HRA ethical approval, which was reviewed and approved by South Yorkshire Research Ethics Committee (REC No.20/YH/0121). We also accessed 1000 anonymous residual blood samples from NHSBT convalescent plasma donated between 19^th^ May and 26^th^ June.

### Serology

Red cells were tested serologically for ABO, Rh and Lewis using relevant DiaClon cards in accordance with the manufacturer’s instructions (Bio-Rad).

### Genomic DNA isolation

Genomic DNA was isolated from whole blood samples using the PureLink Genomic DNA Mini kit according to the manufacturer’s instructions (Invitrogen).

### Genotyping of the FUT2 G428A polymorphism

ABH antigen secretor status was determined by allele-specific PCR of the G428A polymorphism in the FUT2 gene. PCR products of 131-132 bp were obtained using modified versions of primers described by^16^. Detection of the wild-type allele used a G-specific forward primer 5’-CCGGCTACCCCTGCTCGTG-3’ and the common reverse primer 5’-CCGGCTCCCGTTCACCTG-3’. Detection of the null allele that prevents secretion used an A-specific forward primer 5’-ACCGGCTACCCCTGCTCGTA-3’ with the common reverse primer. Samples with discrepant results for genotyping and Lewis serology underwent sequencing of the *FUT2* and *FUT3* coding regions using primers described by King et al 2019.

### ABO genotyping by allelic discrimination

ABO genotype of the DISCOVER DNA samples was determined using three allelic discrimination assays to assess the polymorphisms at positions 261 (+/-G), 526 (C/G) and 703 (G/A) of the ABO gene. The assay for position 526 was the TaqMan SNP genotyping assay C__27859399_10 (SNP ID rs7853989) (ThermoFisher Scientific). The other two assays used primers and TaqMan probes designed by Molecular Diagnostics, NHSBT. Sequences are available on request from the authors. All assays were run in 20ul volumes on a real-time PCR system, according to manufacturer’s instructions (Applied Biosystems).

### Statistics

Statistical analysis were performed with the use of R (v4.0.0)^17^ and respective “pubh” package (v1.3.2)^18^. Statistical comparisons for discrete variables were performed using the two-tailed Fisher’s exact test, and statistical comparisons for continuous variables were performed with the Wilcoxon rank sum test using blood group O as the baseline for comparison. Where necessary, ABO and secretor frequencies were compared using Pearson’s chi-squared test against frequencies reported in the official statistics provided by NHS Blood and Transplant, comprising blood group distribution in the England.

## Results

### COVID-19 patients with Blood Group A are more likely to be hospitalized and suffer cardiovascular complications

A total of 471 adult patients had been admitted to North Bristol NHS Trust (UK) with a positive PCR result for SARS-CoV-2 and ABO blood group data was available for 44% (n=209) of these. Retrospective analysis of these data revealed that among all the blood types, blood type A was the most common in COVID-19 patients, followed by O, B and then type AB as the least common (**Table 1** and **Supplemental Table 1**). As observed in other similar studies, the proportion of blood group A in patients with COVID-19 was significantly higher than those with blood group O; with 105 (50.2%) patients with type A blood and 83 (39.7%) with type O (RR=1.27, CI 95% [1.03, 1.55], P=0.0246) (Table 2). By comparison, in the English donor population type O is the most common blood group accounting for 48% of the population while type A accounts for 38% of the population.^19^ The increased risk of hospitalisation for blood group A for COVID-19 is significant and is accompanied by a significantly higher instance of respiratory failure on admission, requiring ventilation (P=0.01525). However, there was no significant association with length of stay in hospital or patient death rate. Hospitalised type A and O COVID-19 patients also had a similar age range (average +/- SD for A = 74.5 +/- 16.9 and O=72.6 +/- 17.9). Interestingly, we observed a striking gender distribution in this study (**Table 1 and Table 2**) with 64% of males in the type A group compared to 43% with O type and conversely, more females with type O compared to A (56% vs 35%) in this cohort. These observations match previous reports that male gender as a risk factor for COVID-19 ^20^, but this variation is unexplained in the context of blood type.

**Table 1.** Retrospective analysis of critically ill patients admitted to the intensive care in North Bristol NHS Trust (UK) with a positive PCR result for SARS-CoV-2 and that for whom ABO blood group data was available.

|  |  | ABO blood group |  |  |  |  |
| --- | --- | --- | --- | --- | --- | --- |
|  |  | A | AB | B | O | Total |
| Number / Percentage of total |  | 105 (50.24%) | 3 (1.44%) | 18 (8.61%) | 83 (39.71%) | 209 (100%) |
| Age (± SD, years) |  | 74.5 ± 16.9 | 87.8 ± 2.8 | 68.3 ± 17.6 | 72.6 ± 17.9 | 73.4 ± 17.3 |
| Gender |  |  |  |  |  |  |
| Female |  | 37 (35.2%) | 0 (0.0%) | 11 (61.1%) | 47 (56.6%) | 95 (45.5%) |
| Male |  | 68 (64.8%) | 3 (100.0%) | 7 (38.9%) | 36 (43.4%) | 114 (54.5%) |
| Respiratory failure at admission |  |  |  |  |  |  |
| No |  | 58 (55.2%) | 3 (100.0%) | 11 (61.1%) | 61 (73.5%) | 133 (63.6%) |
| Yes |  | 47 (44.8%) | 0 (0.0%) | 7 (38.9%) | 22 (26.5%) | 76 (36.4%) |
| Length of hospital stay (± SD, days) |  | 16.3 ± 17.6 | 17.0 ± 13.9 | 16.9 ± 25.0 | 16.0 ± 20.4 | 16.2 ± 19.3 |
| Inpatient death |  |  |  |  |  |  |
| Unknown/No data |  | 2 (1.9%) | 0 (0.0%) | 0 (0.0%) | 0 (0.0%) | 2 (1.0%) |
| No |  | 72 (68.6%) | 2 (66.7%) | 15 (83.3%) | 57 (68.7%) | 146 (69.9%) |
| Yes |  | 31 (29.5%) | 1 (33.3%) | 3 (16.7%) | 26 (31.3%) | 61 (29.2%) |
| Acute renal failure |  |  |  |  |  |  |
| No |  | 74 (70.5%) | 1 (33.3%) | 12 (66.7%) | 63 (75.9%) | 150 (71.8%) |
| Yes |  | 31 (29.5%) | 2 (66.7%) | 6 (33.3%) | 20 (24.1%) | 59 (28.2%) |
| Liver dysfunction |  |  |  |  |  |  |
| No |  | 95 (90.5%) | 3 (100.0%) | 16 (88.9%) | 74 (89.2%) | 188 (90.0%) |
| Yes |  | 10 (9.5%) | 0 (0.0%) | 2 (11.1%) | 9 (10.8%) | 21 (10.0%) |
| ARDS (Acute respiratory distress syndrome) |  |  |  |  |  |  |
| No |  | 94 (89.5%) | 3 (100.0%) | 14 (77.8%) | 76 (91.6%) | 187 (89.5%) |
| Yes |  | 11 (10.5%) | 0 (0.0%) | 4 (22.2%) | 7 (8.4%) | 22 (10.5%) |
| Cardiovascular complication* |  |  |  |  |  |  |
| No |  | 75 (71.4%) | 3 (100.0%) | 16 (88.9%) | 70 (84.3%) | 164 (78.5%) |
| Yes |  | 30 (28.6%) | 0 (0.0%) | 2 (11.1%) | 13 (15.7%) | 45 (21.5%) |
| No complications |  |  |  |  |  |  |
| No |  | 77 (73.3%) | 2 (66.7%) | 9 (50.0%) | 48 (57.8%) | 136 (65.1%) |
| Yes |  | 28 (26.7%) | 1 (33.3%) | 9 (50.0%) | 35 (42.2%) | 73 (34.9%) |

\*Cardiovascular complications encompasses non-ST-elevation myocardial infarction (NSTEMI), ST-elevation myocardial infarction (STEMI), new episode atrial fibrillation, stroke or brain haemorrhage, deep vein thrombus (DVT), pulmonary embolus (PE) and new/worsening congestive heart failure.

**Table 2.**
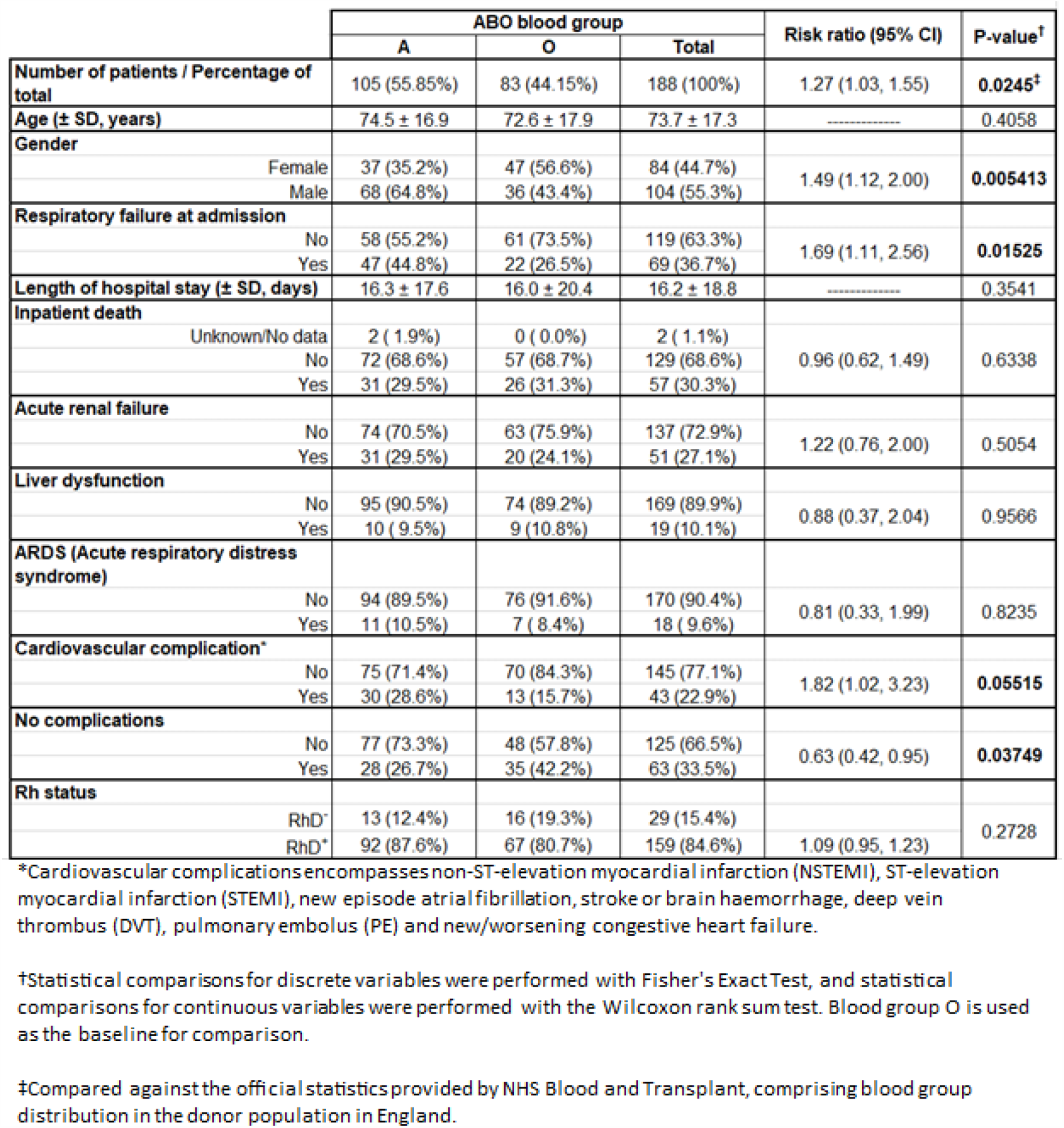
Statistical analysis of data produced from Table 1.

Importantly, we observe a trend between blood group A status and complications with cardiovascular disease (**Table 1, Table 2** and **Supplemental Table 1**). Individuals with blood group type A displayed almost double the risk of suffering from a cardiovascular complication compared to individuals with type O (RR = 1.82, CI 95% [1.02, 3.23], P=0.055). In contrast, there was no observed association between A and O blood group and suffering from acute respiratory distress syndrome (ARDS, RR= 0.81, CI 95% [0.33,1.99], P=0.8235). As a control comparison, we did not detect a significant difference in the frequency of RhD phenotype between A and O (RR= 1.09, CI 95% [0.95, 1.23], P= 0.273). Further statistical analysis was performed by dividing A and O type populations into two subgroups by gender (**Table 3**). Whilst the risk ratios remained supportive of the hypothesis that the A blood group is associated with cardiovascular complications in COVID-19 patients regardless of gender (Male: RR=1.59, CI 95% [0.75,3.33], P=0.3083; Female: RR=1.92, CI 95% [0.75, 4.76], P=0.2774), the two comparisons were not statistically meaningful, likely due to the decrease in number of patients per compared group.

**Table 3.**
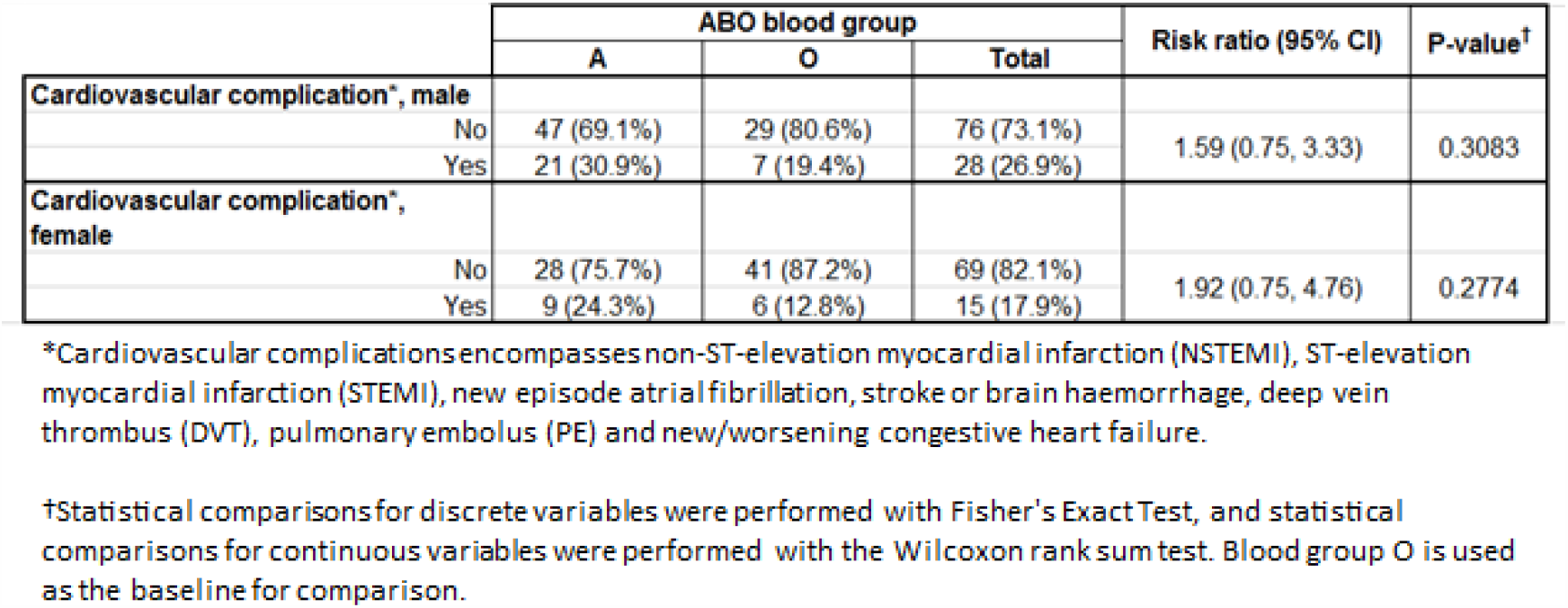
Statistical analysis of patients with cardiovascular complications, divided on gender, from Table 1.

### Secretor status is a compounding risk factor for hospitalization of group A COVID-19 patients

To further investigate the association of COVID-19 with blood group type, blood samples collected by the DISCOVER study were tested for ABO group and secretor status using DNA-based methodologies (**Table 4**. See **Supplemental Table 2** for clinical data on admission). Whilst the two cohorts have different inclusion criteria, a retrospective analysis showed 24 patients were included in both cohorts, and these comprised approximately 10% of the Avon CAP cohort and 20% of the DISCOVER cohort. Again, we observed a similar trend for the association of blood group A with cardiovascular complications in COVID-19 as compared to O which is significant (RR = 2.00, CI 95% [1.01, 3.95], P=0.074). When the laboratory blood group data from non-overlapping patients confirmed PCR positive for SARS-CoV2 data were combined with the earlier health surveillance data, (n=275) the association of blood group A with hospitalization (RR=1.24, CI 95% [1.05,1.47], P=0.0111) and blood group A with cardiovascular complications (RR=2.56, CI 95% [1.43,4.55], P=0.0011) now have increased significance. (see also **Supplemental Table 3 and 4**). The blood group A were associated with cardiovascular complications in COVID-19 patients regardless of gender and was significant with the larger group of patients (Male: RR=2.50, CI 95% [1.18,5.26], P=0.0168; Female: RR=2.33, CI 95% [0.95, 5.88], P=0.0919)

**Table 4.** Retrospective analysis of patients admitted to North Bristol NHS Trust (UK) and enrolled onto the DISCOVER study, that were phenotyped and genotyped for ABO blood group and for secretor status. Highlighted in bold are cardiovascular complications as a result of COVID19 infection. Cardiovascular complications are classed as patients requiring or developing inotropic support, NSTEMI, STEMI, myocarditis, new episode of atrial fibrillation, new or worsening congestive heart failure or new DVT/PE.

|  |  | ABO Blood groups |  |  |  |  | Secretor phenotype |  |  |
| --- | --- | --- | --- | --- | --- | --- | --- | --- | --- |
|  |  | A<br>(N=62) | AB<br>(N=8) | B<br>(N=20) | O<br>(N=53) | Total<br>(N=143) | Non-Secretor<br>(N=26) | Secretor<br>(N=117) | Total<br>(N=143) |
| ABO | A |  |  |  |  |  | 5 (19.2%) | 57 (48.7%) | 62 (43.4%) |
|  | AB |  |  |  |  |  | 3 (11.5%) | 5 (4.3%) | 8 (5.6%) |
|  | B |  |  |  |  |  | 5 (19.2%) | 15 (12.8%) | 20 (14.0%) |
|  | O |  |  |  |  |  | 13 (50.0%) | 40 (34.2%) | 53 (37.1%) |
| PCR Secretor | Non-Secretor | 5 (8.1%) | 3 (37.5%) | 5 (25.0%) | 13 (24.5%) | 26 (18.2%) |  |  |  |
|  | Secretor | 57 (91.9%) | 5 (62.5%) | 15 (75.0%) | 40 (75.5%) | 117 (81.8%) |  |  |  |
| Age |  | 60.5 ± 18.3 | 63.6 ± 17.4 | 52.8 ± 11.4 | 58.1 ± 14.5 | 58.7 ± 16.1 | 58.0 ± 12.7 | 58.9 ± 16.8 | 58.7 ± 16.1 |
| Sex | Female | 27 (43.5%) | 2 (25.0%) | 8 (40.0%) | 21 (39.6%) | 58 (40.6%) | 13 (50.0%) | 45 (38.5%) | 58 (40.6%) |
|  | Male | 35 (56.5%) | 6 (75.0%) | 12 (60.0%) | 32 (60.4%) | 85 (59.4%) | 13 (50.0%) | 72 (61.5%) | 85 (59.4%) |
| Caucasian | No | 9 (14.5%) | 1 (12.5%) | 5 (25.0%) | 6 (11.3%) | 21 (14.7%) | 5 (19.2%) | 16 (13.7%) | 21 (14.7%) |
|  | Unknown | 6 (9.7%) | 3 (37.5%) | 3 (15.0%) | 5 (9.4%) | 17 (11.9%) | 6 (23.1%) | 11 (9.4%) | 17 (11.9%) |
|  | Yes | 47 (75.8%) | 4 (50.0%) | 12 (60.0%) | 42 (79.2%) | 105 (73.4%) | 15 (57.7%) | 90 (76.9%) | 105 (73.4%) |
| Preconditions Diabetes | No | 47 (75.8%) | 7 (87.5%) | 16 (80.0%) | 43 (81.1%) | 113 (79.0%) | 22 (84.6%) | 91 (77.8%) | 113 (79.0%) |
|  | T1DM | 2 (3.2%) | 0 (0.0%) | 0 (0.0%) | 1 (1.9%) | 3 (2.1%) | 0 (0.0%) | 3 (2.6%) | 3 (2.1%) |
|  | T2DM | 13 (21.0%) | 1 (12.5%) | 4 (20.0%) | 9 (17.0%) | 27 (18.9%) | 4 (15.4%) | 23 (19.7%) | 27 (18.9%) |
| Preconditions Heart disease | No | 42 (67.7%) | 6 (75.0%) | 17 (85.0%) | 44 (83.0%) | 109 (76.2%) | 20 (76.9%) | 89 (76.1%) | 109 (76.2%) |
|  | Yes | 20 (32.3%) | 2 (25.0%) | 3 (15.0%) | 9 (17.0%) | 34 (23.8%) | 6 (23.1%) | 28 (23.9%) | 34 (23.8%) |
| Preconditions Hypertension | No | 41 (66.1%) | 7 (87.5%) | 15 (75.0%) | 38 (71.7%) | 101 (70.6%) | 19 (73.1%) | 82 (70.1%) | 101 (70.6%) |
|  | Yes | 21 (33.9%) | 1 (12.5%) | 5 (25.0%) | 15 (28.3%) | 42 (29.4%) | 7 (26.9%) | 35 (29.9%) | 42 (29.4%) |
| Preconditions Chronic Lung disease | No | 43 (69.4%) | 6 (75.0%) | 12 (60.0%) | 44 (83.0%) | 105 (73.4%) | 18 (69.2%) | 87 (74.4%) | 105 (73.4%) |
|  | Yes | 19 (30.6%) | 2 (25.0%) | 8 (40.0%) | 9 (17.0%) | 38 (26.6%) | 8 (30.8%) | 30 (25.6%) | 38 (26.6%) |
| Preconditions Severe Liver disease | No | 61 (98.4%) | 8 (100.0%) | 20 (100.0%) | 52 (98.1%) | 141 (98.6%) | 26 (100.0%) | 115 (98.3%) | 141 (98.6%) |
|  | Yes | 1 (1.6%) | 0 (0.0%) | 0 (0.0%) | 1 (1.9%) | 2 (1.4%) | 0 (0.0%) | 2 (1.7%) | 2 (1.4%) |
| Preconditions Severe kidney impairment | No | 58 (93.5%) | 7 (87.5%) | 20 (100.0%) | 44 (83.0%) | 129 (90.2%) | 25 (96.2%) | 104 (88.9%) | 129 (90.2%) |
|  | Yes | 4 (6.5%) | 1 (12.5%) | 0 (0.0%) | 9 (17.0%) | 14 (9.8%) | 1 (3.8%) | 13 (11.1%) | 14 (9.8%) |
| Symptoms Respiratory | No | 11 (17.7%) | 0 (0.0%) | 0 (0.0%) | 6 (11.3%) | 17 (11.9%) | 0 (0.0%) | 17 (14.5%) | 17 (11.9%) |
|  | Yes | 51 (82.3%) | 8 (100.0%) | 20 (100.0%) | 47 (88.7%) | 126 (88.1%) | 26 (100.0%) | 100 (85.5%) | 126 (88.1%) |
| Symptoms Systemic | No | 20 (32.3%) | 1 (12.5%) | 1 (5.0%) | 5 (9.4%) | 27 (18.9%) | 3 (11.5%) | 24 (20.5%) | 27 (18.9%) |
|  | Yes | 42 (67.7%) | 7 (87.5%) | 19 (95.0%) | 48 (90.6%) | 116 (81.1%) | 23 (88.5%) | 93 (79.5%) | 116 (81.1%) |
| Symptoms Neurological | No | 54 (87.1%) | 6 (75.0%) | 18 (90.0%) | 47 (88.7%) | 125 (87.4%) | 22 (84.6%) | 103 (88.0%) | 125 (87.4%) |
|  | Yes | 8 (12.9%) | 2 (25.0%) | 2 (10.0%) | 6 (11.3%) | 18 (12.6%) | 4 (15.4%) | 14 (12.0%) | 18 (12.6%) |
| General outcome | Deceased | 5 (8.1%) | 1 (12.5%) | 2 (10.0%) | 3 (5.7%) | 11 (7.7%) | 2 (7.7%) | 9 (7.7%) | 11 (7.7%) |
|  | Discharged | 52 (83.9%) | 7 (87.5%) | 15 (75.0%) | 45 (84.9%) | 119 (83.2%) | 21 (80.8%) | 98 (83.8%) | 119 (83.2%) |
|  | Inpatient | 3 (4.8%) | 0 (0.0%) | 0 (0.0%) | 2 (3.8%) | 5 (3.5%) | 0 (0.0%) | 5 (4.3%) | 5 (3.5%) |
|  | Unknown | 2 (3.2%) | 0 (0.0%) | 3 (15.0%) | 3 (5.7%) | 8 (5.6%) | 3 (11.5%) | 5 (4.3%) | 8 (5.6%) |
| Complications Cardiovascular | No | 49 (79.0%) | 8 (100.0%) | 19 (95.0%) | 52 (98.1%) | 128 (89.5%) | 223 (92.3%) | 104 (88.9%) | 128 (89.5%) |
|  | Yes | 13 (21.0%) | 0 (0.0%) | 1 (5.0%) | 1 (1.9%) | 15 (10.5%) | 3 (7.7%) | 13 (11.1%) | 15 (10.5%) |
| Complications Acute renal failure | No | 57 (91.9%) | 8 (100.0%) | 18 (90.0%) | 46 (86.8%) | 129 (90.2%) | 23 (88.5%) | 106 (90.6%) | 129 (90.2%) |
|  | Yes | 5 (8.1%) | 0 (0.0%) | 2 (10.0%) | 7 (13.2%) | 14 (9.8%) | 3 (11.5%) | 11 (9.4%) | 14 (9.8%) |
| Complications Liver dysfunction | No | 50 (80.6%) | 7 (87.5%) | 19 (95.0%) | 46 (86.8%) | 122 (85.3%) | 22 (84.6%) | 100 (85.5%) | 122 (85.3%) |
|  | Yes | 12 (19.4%) | 1 (12.5%) | 1 (5.0%) | 7 (13.2%) | 21 (14.7%) | 4 (15.4%) | 17 (14.5%) | 21 (14.7%) |

In epithelial tissues and secretions, ABO expression is heavily dependent on the inheritance of the Secretor *Se/FUT2* gene which can also be protective against viral infection. Due to mutations in other fucosyltransferase genes individuals can also be Le^a-b-^, and rarely Le^a+b+^ so secretor status cannot always be determined by red cell typing alone. We therefore conducted DNA analysis on this cohort to further determine ABO and secretor status and subsequently compared all symptom and lab values to ABO and secretor genotype. Individuals can either be secretors (SeSe or Sese) or non-secretors (sese). Strikingly, we observed that the vast majority of blood group A patients expressed an active Secretor gene (*FUT2*), with the incidence of non-secretors being significantly lower than would be expected from comparison with the normal distribution in the general population (**Table 4**, (8.1% vs. 20%, P = 0.019).^21^ No initial correlation with Se genotype (SeSe versus Sese) and disease outcome was observed. After additional blood group genotype analysis this showed there were 3 deaths amongst only 9 AA Se/Se or Se/se patients compared with 2 deaths from 53 AO Se/Se or Se/se patients (**Table 5**) but we caution any extrapolation from this result as the sample size is small.

**Table 5.** Retrospective analysis of patients admitted to North Bristol NHS Trust (UK) and enrolled onto the DISCOVER study, that were genotyped for ABO blood group and secretor status. Highlighted in bold are cardiovascular complications as a result of COVID19 infection. Cardiovascular complications are classed as patients requiring or developing inotropic support, NSTEMI, STEMI, myocarditis, new episode of atrial fibrillation, new or worsening congestive heart failure or new DVT/PE.

|  |  | ABO genotype |  |  |  |  |  |  | Secretor genotype |  |  |  |
| --- | --- | --- | --- | --- | --- | --- | --- | --- | --- | --- | --- | --- |
|  |  | AA<br>(N=9) | AO<br>(N=53) | AB<br>(N=8) | BB<br>(N=1) | BO<br>(N=18) | OO<br>(N=53) | Total<br>(N=142) | sese<br>(N=25) | SeSe<br>(N=47) | Sese<br>(N=70) | Total<br>(N=142) |
| ABO | AA |  |  |  |  |  |  |  | 1 (4.0%) | 0 (0.0%) | 8 (11.4%) | 9 (6.3%) |
|  | AO |  |  |  |  |  |  |  | 4 (16.0%) | 18 (38.3%) | 31 (44.3%) | 53 (37.3%) |
|  | AB |  |  |  |  |  |  |  | 3 (12.0%) | 0 (0.0%) | 5 (7.1%) | 8 (5.6%) |
|  | BB |  |  |  |  |  |  |  | 0 (0.0%) | 0 (0.0%) | 1 (1.4%) | 1 (0.7%) |
|  | BO |  |  |  |  |  |  |  | 4 (16.0%) | 9 (19.1%) | 5 (7.1%) | 18 (12.7%) |
|  | OO |  |  |  |  |  |  |  | 13 (52.0%) | 20 (42.6%) | 20 (28.6%) | 53 (37.3%) |
| PCR Secretor |  |  |  |  |  |  |  |  |  |  |  |  |
| sese (Non-Secretor) |  | 1 (11.2%) | 4 (7.5%) | 3 (37.5%) | 0 (0.0%) | 4 (22.2%) | 13 (24.5%) | 25 (17.6%) |  |  |  |  |
| Sese (Secretor) |  | 8 (88.8%) | 31 (58.5%) | 5 (62.5%) | 1 (100%) | 5 (27.8%) | 20 (37.7%) | 70 (49.3%) |  |  |  |  |
| SeSe (Secretor) |  | 0 (0.0%) | 18 (34.0%) | 0 (0.0%) | 0 (0.0%) | 9 (50.0%) | 20 (37.7%) | 47 (33.1%) |  |  |  |  |
| Age |  | 62.9 ± 26.9 | 59.9 ± 39.9 | 63.6 ± 17.4 | 31 ± 0 | 54.4 ± 12.4 | 58.1 ± 14.5 | 58.7 ± 16.1 | 58.1 ± 12.7 | 61.6 ± 31.3 | 57.1 ± 30.9 | 58.7 ± 16.1 |
| Sex |  |  |  |  |  |  |  |  |  |  |  |  |
| Female |  | 2 (22.2%) | 25 (47.2%) | 2 (25.0%) | 0 (0.0%) | 7 (38.9%) | 21 (39.6%) | 57 (40.1%) | 12 (48.0%) | 20 (42.6%) | 25 (35.7%) | 57 (40.1%) |
| Male |  | 7 (77.8%) | 28 (52.8%) | 6 (75.0%) | 1 (100%) | 11 (61.1%) | 32 (60.4%) | 85 (59.9%) | 13 (52.0%) | 27 (57.4%) | 45 (64.3%) | 85 (59.9%) |
| Caucasian |  |  |  |  |  |  |  |  |  |  |  |  |
| No |  | 0 (0.0%) | 9 (17.0%) | 1 (12.5%) | 0 (0.0%) | 4 (22.2%) | 42 (79.2%) | 56 (39.4%) | 5 (20.0%) | 7 (14.9%) | 8 (11.4%) | 20 (14.1%) |
| Unknown |  | 2 (22.2%) | 4 (7.5%) | 3 (37.5%) | 0 (0.0%) | 3 (16.7%) | 5 (9.4%) | 17 (12.0%) | 5 (20.0%) | 6 (12.8%) | 6 (8.6%) | 17 (12.0%) |
| Yes |  | 7 (77.8%) | 40 (75.5%) | 4 (50.0%) | 1 (100%) | 11 (61.1%) | 6 (11.3%) | 69 (48.6%) | 15 (60.0%) | 34 (72.3%) | 56 (80.0%) | 105 (73.9%) |
| Preconditions Diabetes |  |  |  |  |  |  |  |  |  |  |  |  |
| No |  | 8 (88.8%) | 39 (73.6%) | 7 (87.5%) | 1 (100%) | 14 (77.8%) | 43 (81.1%) | 112 (78.9%) | 21 (84.0%) | 33 (70.2%) | 58 (82.9%) | 112 (78.9%) |
| T1DM |  | 0 (0.0%) | 2 (3.8%) | 0 (0.0%) | 0 (0.0%) | 0 (0.0%) | 1 (1.9%) | 3 (2.1%) | 0 (0.0%) | 1 (2.1%) | 2 (2.9%) | 3 (2.1%) |
| T2DM |  | 1 (11.2%) | 12 (22.6%) | 1 (12.5%) | 0 (0.0%) | 4 (22.2%) | 9 (17.0%) | 27 (19.0%) | 4 (16.0%) | 13 (27.7%) | 10 (14.3%) | 27 (19.0%) |
| Preconditions Heart disease |  |  |  |  |  |  |  |  |  |  |  |  |
| No |  | 5 (55.5%) | 37 (70.0%) | 6 (75.0%) | 1 (100%) | 15 (83.3%) | 44 (83.0%) | 108 (76.1%) | 20 (80.0%) | 38 (80.9%) | 51 (71.9%) | 109 (76.8%) |
| Yes |  | 4 (44.5%) | 16 (30%) | 2 (25.0%) | 0 (0.0%) | 3 (16.7%) | 9 (17.0%) | 34 (23.9%) | 5 (20.0%) | 9 (19.1) | 19 (27.1%) | 33 (23.2%) |
| Preconditions Hypertension |  |  |  |  |  |  |  |  |  |  |  |  |
| No |  | 5 (55.5%) | 36 (67.9%) | 7 (87.5%) | 1 (100%) | 13 (72.2%) | 38 (71.7%) | 100 (70.4%) | 18 (72.0%) | 32 (68.1%) | 50 (71.4%) | 100 (70.4%) |
| Yes |  | 4 (44.5%) | 17 (32.1%) | 1 (12.5%) | 0 (0.0%) | 5 (27.8%) | 15 (28.3%) | 42 (29.6%) | 7 (28.0%) | 15 (31.9%) | 20 (28.6%) | 42 (29.6%) |
| Preconditions Chronic Lung disease |  |  |  |  |  |  |  |  |  |  |  |  |
| No |  | 7 (77.8%) | 36 (67.9%) | 6 (75.0%) | 1 (100%) | 10 (55.6%) | 44 (83.0%) | 104 (73.2%) | 17 (68.0%) | 33 (70.2%) | 54 (77.1%) | 104 (73.2%) |
| Yes |  | 2 (22.2%) | 17 (32.1%) | 2 (25.0%) | 0 (0.0%) | 8 (44.4%) | 9 (17.0%) | 38 (26.8%) | 8 (32.0%) | 14 (29.8%) | 16 (22.9%) | 38 (26.8%) |
| Preconditions Severe Liver disease |  |  |  |  |  |  |  |  |  |  |  |  |
| No |  | 9 (100%) | 52 (98.2%) | 8 (100%) | 1 (100%) | 18 (100%) | 52 (98.1%) | 140 (98.6%) | 25 (100%) | 47 (100%) | 68 (97.1%) | 140 (98.6%) |
| Yes |  | 0 (0.0%) | 1 (1.8%) | 0 (0.0%) | 0 (0.0%) | 0 (0.0%) | 1 (1.9%) | 2 (1.4%) | 0 (0.0%) | 0 (0.0%) | 2 (2.9%) | 2 (1.4%) |
| Preconditions Severe kidney impairment |  |  |  |  |  |  |  |  |  |  |  |  |
| No |  | 8 (88.8%) | 50 (94.3%) | 7 (87.5%) | 1 (100%) | 18 (100%) | 44 (83.0%) | 128 (90.1%) | 24 (96.0%) | 41 (87.2%) | 63 (90.0%) | 128 (90.1%) |
| Yes |  | 1 (11.2%) | 3 (5.7%) | 1 (12.5%) | 0 (0.0%) | 0 (0.0%) | 9 (17.0%) | 14 (9.9%) | 1 (4.0%) | 6 (12.8%) | 7 (10.0%) | 14 (9.9%) |
| Symptoms Respiratory |  |  |  |  |  |  |  |  |  |  |  |  |
| No |  | 2 (22.2%) | 9 (17.0%) | 0 (0.0%) | 0 (0.0%) | 18 (100%) | 6 (11.3%) | 35 (24.6%) | 0 (0.0%) | 5 (10.6%) | 12 (17.1%) | 17 (12.0%) |
| Yes |  | 7 (77.7%) | 44 (83.0%) | 8 (100%) | 1 (100%) | 0 (0.0%) | 47 (88.7%) | 107 (75.4%) | 25 (100%) | 42 (89.4%) | 58 (82.9%) | 125 (88.0%) |
| Symptoms Systemic |  |  |  |  |  |  |  |  |  |  |  |  |
| No |  | 2 (22.2%) | 18 (34.0%) | 1 (12.5%) | 0 (0.0%) | 1 (5.6%) | 5 (9.4%) | 27 (19%) | 2 (8.0%) | 10 (21.3%) | 14 (20.0%) | 26 (18.3%) |
| Yes |  | 7 (77.8%) | 35 (66.0%) | 7 (87.5%) | 1 (100%) | 17 (94.4%) | 48 (90.6%) | 115 (81%) | 23 (92.0%) | 37 (78.7%) | 56 (80.0%) | 116 (81.7%) |
| Symptoms Neurological |  |  |  |  |  |  |  |  |  |  |  |  |
| No |  | 8 (88.8%) | 46 (86.8%) | 6 (75.0%) | 1 (100%) | 16 (88.9%) | 47 (88.7%) | 124 (87.3%) | 21 (84.0%) | 40 (85.1%) | 63 (90.0%) | 124 (87.4%) |
| Yes |  | 1 (11.2%) | 7 (13.2%) | 2 (25.0%) | 0 (0.0%) | 2 (11.1%) | 6 (11.3%) | 18 (12.7%) | 4 (16.0%) | 7 (14.9%) | 7 (10.0%) | 18 (12.7%) |
| General outcome |  |  |  |  |  |  |  |  |  |  |  |  |
| Deceased |  | 3 (33.3%) | 2 (3.8%) | 0 (0.0%) | 0 (0.0%) | 2 (11.1%) | 3 (5.7%) | 10 (7.0%) | 1 (4.0%) | 2 (4.3%) | 7 (10.0%) | 10 (7.0%) |
| Discharged |  | 5 (55.5%) | 47 (88.7%) | 7 (87.5%) | 1 (100%) | 13 (72.2%) | 45 (84.9%) | 118 (83.1%) | 21 (84.0%) | 40 (85.1%) | 57 (81.4%) | 118 (83.1%) |
| Inpatient |  | 0 (0.0%) | 3 (5.6%) | 0 (0.0%) | 0 (0.0%) | 0 (0.0%) | 2 (3.8%) | 5 (3.5%) | 0 (0.0%) | 0 (0.0%) | 4 (5.7%) | 4 (2.8%) |
| Unknown |  | 1 (11.2%) | 1 (1.8%) | 1 (12.5%) | 0 (0.0%) | 3 (16.7%) | 3 (5.7%) | 9 (6.3%) | 3 (12.0%) | 5 (10.6%) | 2 (2.9%) | 10 (7.0%) |
| Complications Cardiovascular |  |  |  |  |  |  |  |  |  |  |  |  |
| No |  | 7 (77.8%) | 42 (77.3%) | 8 (100%) | 1 (100%) | 17 (94.4%) | 52 (98.1%) | 127 (89.4%) | 22 (88.0%) | 42 (89.4%) | 62 (88.6%) | 126 (88.7%) |
| Yes |  | 2 (22.2%) | 11 (20.7%) | 0 (0.0%) | 0 (0.0%) | 1 (5.6%) | 1 (1.9%) | 15 (10.6%) | 3 (12.0%) | 5 (10.6%) | 8 (11.4%) | 16 (11.3%) |
| Complications Acute renal failure |  |  |  |  |  |  |  |  |  |  |  |  |
| No |  | 8 (88.8%) | 49 (92.4%) | 8 (100%) | 1 (100%) | 16 (88.9%) | 46 (86.8%) | 128 (90.1%) | 22 (88.0%) | 41 (87.2%) | 65 (92.9%) | 128 (90.1%) |
| Yes |  | 1 (11.2%) | 4 (7.5%) | 0 (0.0%) | 0 (0.0%) | 2 (11.1%) | 7 (13.2%) | 14 (9.9%) | 3 (12.0%) | 6 (12.8%) | 5 (7.1%) | 14 (9.9%) |
| Complications Liver dysfunction |  |  |  |  |  |  |  |  |  |  |  |  |
| No |  | 8 (88.8%) | 42 (79.2%) | 7 (87.5%) | 1 (100%) | 17 (94.4%) | 46 (86.8%) | 121 (86.2%) | 21 (84.0%) | 41 (87.2%) | 59 (84.3%) | 121 (85.2%) |
| Yes |  | 1 (11.2%) | 11 (20.8%) | 1 (12.5%) | 0 (0.0%) | 1 (5.6%) | 7 (13.2%) | 21 (14.8%) | 4 (16.0%) | 6 (12.8%) | 11 (15.7%) | 21 (14.8%) |

In order to determine whether non-secretor status among group A individuals was associated with increased protection against SARS-CoV-2 or whether it simply reduced disease severity, residual testing samples were accessed from the UK wide NHSBT convalescent plasma donations collected from recovered COVID-19 patients^22^. The convalescent plasma samples include a broader range of donors who had self-reported recovery from hospitalisation, donors who had positive PCR tests and donors who had suffered known symptoms and undergone a positive antibody test. In confirmation of our previous results, we observed a lower than expected number of non-secretors in blood group A donors who had reported hospitalisation (N= 55), but note that this analysis is based on a small sample size (**Table 6**). Importantly, across all convalescent donors sampled we observed the anticipated number of non-secretors. However, in patients with blood type A, a higher-than-expected number of non-secretors was identified (25% vs 20%, P = 0.01) (**Table 6**). Taken together, the DISCOVER study and convalescent plasma donor study results suggest that blood type A non-secretors are not necessarily protected from SARS-CoV-2 infection but may experience less severe disease.

**Table 6.** ABO and Secretor phenotype of patients admitted to North Bristol NHS Trust (UK) and enrolled onto the DISCOVER study, samples from known hospitalised COVID19 NHSBT convalescent plasma donations and a much larger cohort of non-hospitalised NHSBT convalescent plasma donations. Highlighted in bold are the numbers and percentages of secretors and non-secretors of blood type A in each cohort.

| DISCOVER |  |  |  |  |  |  |
| --- | --- | --- | --- | --- | --- | --- |
|  |  | A<br>(N=62) | AB<br>(N=8) | B<br>(N=20) | O<br>(N=53) | Total<br>(N=143) |
| PCR Secretor |  |  |  |  |  |  |
| Non-Secretor |  | 5 (8.1%) | 3 (37.5%) | 5 (25.0%) | 13 (24.5%) | 26 (18.2%) |
| Secretor |  | 57 (91.9%) | 5 (62.5%) | 15 (75.0%) | 40 (75.5%) | 117 (81.8%) |

| Hospitalised Convalescent Plasma |  |  |  |  |  |  |
| --- | --- | --- | --- | --- | --- | --- |
|  |  | A<br>(N=23) | AB<br>(N=4) | B<br>(N=6) | O<br>(N=22) | Total<br>(N=55) |
| PCR Secretor |  |  |  |  |  |  |
| Non-Secretor |  | 3 (13.0%) | 1 (25.0%) | 2 (33.3%) | 5 (22.7%) | 11 (20.0%) |
| Secretor |  | 20 (87.0%) | 3 (75.0%) | 4 (66.6%) | 17 (77.3%) | 44 (80.0%) |

**Table 6** – ABO and Secretor phenotype of patients admitted to North Bristol NHS Trust (UK) and enrolled onto the DISCOVER study, samples from known hospitalised COVID19 NHSBT convalescent plasma donations and a much larger cohort of non-hospitalised NHSBT convalescent plasma donations.
| Non-Hospitalised Convalescent Plasma |  |  |  |  |  |  |
| --- | --- | --- | --- | --- | --- | --- |
|  |  | A<br>(N=392) | AB<br>(N=42) | B<br>(N=97) | O<br>(N=397) | Total<br>(N=928) |
| PCR Secretor |  |  |  |  |  |  |
| Non-Secretor |  | 101 (25.8%) | 8 (19.0%) | 19 (19.6%) | 93 (23.4%) | 221 (23.8%) |
| Secretor |  | 291 (74.2%) | 34 (81.0%) | 78 (80.4%) | 304 (76.6%) | 707 (76.2%) |

## Discussion

Studies carried out in hospitalised SARS-CoV-2 patients in China were the first to link blood group A with greater susceptibility to COVID-19 compared to blood group O^15^. Since then, multiple other studies carried out in other countries have supported this conclusion^23-26^. The reasons for the association of severe COVID-19 with blood type A are unknown, but it has been suggested that this could be caused by O group patients having anti-A type antibodies ^27^, that A type glycans could function as co-receptors for SARS-CoV-2^28^ or due to the known effects of blood groups on thrombosis risk due to von Willebrand factor (VWF) levels.^29^ However, recently, the conclusion that ABO group influences COVID-19 severity has been questioned. ^30,31^

Here we report a significant increased risk of hospitalisation for blood group A COVID19 patients compared to patients with blood group O. This was accompanied by a significantly higher instance of respiratory failure on admission, requiring ventilation but no significant increase in patient death. Our data link blood group A preponderance in COVID-19 with cardiovascular outcomes. We found no association with blood group A and development of ARDS, suggesting that SARS-CoV-2 does not bind preferentially to the A blood group structures, as is the case in several other infectious diseases.^8^ This is result is consistent with studies of younger healthier populations where no bias to blood group A over group O was observed. ^31^ We propose that the apparently conflicting results between studies to date can be explained by the nature of the patient population studied, because many studies focus on patient populations ill enough to need hospital admission. Such individuals are mostly elderly and more likely to have co-morbidities.

The association of blood group A with cardiac disease is well documented and is linked to VWF (reviewed by^32^). Our data are the first to specifically link the severity of disease in COVID-19 patients to homozygous group A individuals who are also secretors. Levels of VWF in plasma are highest in homozygous group A individuals^33^ and homozygous Se individuals.^33^ We propose this is likely linked to the degree of N-glycosylation of VWF particularly at position 1574 near the site of cleavage by ADAMTS13.^34^ It follows that more extensive glycosylation of VWF found in group A secretors but lacking in group O secretors could account for higher circulating levels of VWF in Group A and thereby predispose Group A patients to cardiac problems and thrombosis during infections with SARS-CoV-2. The mechanism whereby infection with SARS-CoV-2 influences this predisposition is currently unknown but COVID-19 has been reported to be associated with inducing a hypercoagulable state.^35^ It is known that VWF protein levels associated with pulmonary vascular endothelial cells are linked to ABO determinants.^36^ We speculate that binding of SARS-CoV-2 virus to its receptor ACE2 in the lungs may activate endothelial cells resulting in enhanced secretion of VWF into peripheral circulation.

These results provide an explanation for discrepant results reported regarding the significance of blood group A and COVID-19 disease severity and indicate that determination of the genotype and secretor status of group A individuals with SARS-CoV-2 infection could be a useful diagnostic aid to the stratification of risk of mortality in hospitalised patients. More extensive studies are needed to further explore the stratification of patients by blood group and thereby facilitate identification of the most at risk to death and to understand the complex disease mechanism that induces the hypercoagulative state.

## Supporting information

Mankelow et al 2020 Supplemental tables

## Data Availability

Data not available due ethical/legal restrictions
Due to the nature of this research, participants of this study did not agree for their data to be shared publicly, so supporting data is not available.
All available data is located in the manuscript and supplemental data

## Acknowledgements

This study was supported by the National Institute for Health Research Blood and Transplant Research Unit (NIHR BTRU) in Red Cell Products (IS-BTU-1214-10032) and the Department of Health (England) (National Health Service Blood and Transplant research and development grant - WP15-04). C.H is funded by a National Institute for Health Research (NIHR) Academic Clinical Fellowship in Respiratory Medicine. The views expressed are those of the authors and not necessarily those of the National Health Service, NIHR, or the Department of Health and Social Care. The DISCOVER study was funded by grants from the Southmead Hospital Charity, and support from the Elizabeth Blackwell Institute, University of Bristol. We wish to thank to Dr Louise Tilley and Terri Stutt (Molecular Diagnostics, NHSBT) for kindly providing reagents and advice for the ABO genotyping assays. We also thank the NHSBT and the NHSBT Convalescent Plasma team for access to residual testing samples. We also thank Alexandra Griffiths for provision of NHSBT Convalescent plasma data (Statistics and Clinical Studies, NHSBT).

## Authorship Contributions

T.J.M processed samples and performed serology and Se genotyping, analysed data, and wrote the paper. B.K.S processed samples and performed serology, Se genotyping and ABO genotyping. P.L.M collated data and performed statistical analysis and wrote the paper. C.J.S, N.M.C, P.G and S.K processed samples and performed serology and Se genotyping. L.N, C.A, J.P, F.M, G.R, F.M, F.H, C.H, A.N and A.C conducted health data surveillance. A.F, D.A, C.H and F.H initiated the patient studies at North Bristol NHS trust, provided clinical information of samples and wrote the paper. D.A, C.H. and A.T instigated the research and wrote the paper.

## References

1. Haldane JBS. Disease and Evolution. Ric Sci Suppl. 1949;19:1045.

2. Szulman AE. The histological distribution of blood group substances A and B in man. J Exp Med. 1960;111:785–800.

3. Kelly RJ, Rouquier S, Giorgi D, Lennon GG, Lowe JB. Sequence and expression of a candidate for the human Secretor blood group alpha(1,2)fucosyltransferase gene (FUT2). Homozygosity for an enzyme-inactivating nonsense mutation commonly correlates with the non-secretor phenotype. J Biol Chem. 1995;270(9):4640–4649.

4. Rouquier S, Lowe JB, Kelly RJ, Fertitta AL, Lennon GG, Giorgi D. Molecular cloning of a human genomic region containing the H blood group alpha(1,2)fucosyltransferase gene and two H locus-related DNA restriction fragments. Isolation of a candidate for the human Secretor blood group locus. J Biol Chem. 1995;270(9):4632–4639.

5. Grubb R. Correlation between Lewis blood group and secretor character in man. Nature. 1948;162(4128):933.

6. Ravn V, Dabelsteen E. Tissue distribution of histo-blood group antigens. APMIS. 2000;108(1):1–28.

7. Anstee DJ. The relationship between blood groups and disease. Blood. 2010;115(23):4635–4643.

8. Taylor SL, McGuckin MA, Wesselingh S, Rogers GB. Infection’s Sweet Tooth: How Glycans Mediate Infection and Disease Susceptibility. Trends Microbiol. 2018;26(2):92–101.

9. Payne DC, Currier RL, Staat MA, et al. Epidemiologic Association Between FUT2 Secretor Status and Severe Rotavirus Gastroenteritis in Children in the United States. JAMA Pediatr. 2015;169(11):1040–1045.

10. Perez-Ortin R, Vila-Vicent S, Carmona-Vicente N, Santiso-Bellon C, Rodriguez-Diaz J, Buesa J. Histo-Blood Group Antigens in Children with Symptomatic Rotavirus Infection. Viruses. 2019;11(4).

11. Nordgren J, Svensson L. Genetic Susceptibility to Human Norovirus Infection: An Update. Viruses. 2019;11(3).

12. Azad MB, Wade KH, Timpson NJ. FUT2 secretor genotype and susceptibility to infections and chronic conditions in the ALSPAC cohort. Wellcome Open Res. 2018;3:65.

13. Zhou P, Yang XL, Wang XG, et al. A pneumonia outbreak associated with a new coronavirus of probable bat origin. Nature. 2020;579(7798):270–273.

14. Wiersinga WJ, Rhodes A, Cheng AC, Peacock SJ, Prescott HC. Pathophysiology, Transmission, Diagnosis, and Treatment of Coronavirus Disease 2019 (COVID-19): A Review. JAMA. 2020;324(8):782–793.

15. Zhao J, Yang Y, Huang H, et al. Relationship between the ABO Blood Group and the COVID-19 Susceptibility. Clin Infect Dis. 2020.

16. Moreno A, Campi C, Escovich L, et al. Analysis of the FUT2 gene and Secretor status in patients with oral lesions. Inmunología. 2009;28:131–134.

17. R: A language and environment for statistical computing. http://www.R-project.org: R Foundation for Statistical Computing; 2014.

18. Athens J. pubh: A Toolbox for Public Health and Epidemiology. R package version. https://CRAN.R-project.org/package=pubh; 2020.

19. NHSBT website. https://www.blood.co.uk/why-give-blood/blood-types/; 2018.

20. Lakbar I, Luque-Paz D, Mege JL, Einav S, Leone M. COVID-19 gender susceptibility and outcomes: A systematic review. PLoS One. 2020;15(11):e0241827.

21. Mourant AE, Kopeć AC, Domaniewska-Sobczak K. The distribution of the human blood groups, and other polymorphisms (ed 2d). London: Oxford University Press; 1976.

22. Roberts DJ, Miflin G, Estcourt L. Convalescent plasma for COVID-19: Back to the future. Transfus Med. 2020;30(3):174–176.

23. Hoiland RL, Fergusson NA, Mitra AR, et al. The association of ABO blood group with indices of disease severity and multiorgan dysfunction in COVID-19. Blood Adv. 2020;4(20):4981–4989.

24. Barnkob MB, Pottegard A, Stovring H, et al. Reduced prevalence of SARS-CoV-2 infection in ABO blood group O. Blood Adv. 2020;4(20):4990–4993.

25. Latz CA, DeCarlo C, Boitano L, et al. Blood type and outcomes in patients with COVID-19. Ann Hematol. 2020;99(9):2113–2118.

26. Leaf RK, Al-Samkari H, Brenner SK, Gupta S, Leaf DE. ABO phenotype and death in critically ill patients with COVID-19. Br J Haematol. 2020;190(4):e204–e208.

27. Yamamoto F, Yamamoto M, Muniz-Diaz E. Blood group ABO polymorphism inhibits SARS-CoV-2 infection and affects COVID-19 progression. Vox Sang. 2020.

28. Breiman A, Ruven-Clouet N, Le Pendu J. Harnessing the natural anti-glycan immune response to limit the transmission of enveloped viruses such as SARS-CoV-2. PLoS Pathog. 2020;16(5):e1008556.

29. O’Sullivan JM, Ward S, Fogarty H, O’Donnell JS. More on ‘Association between ABO blood groups and risk of SARS-CoV-2 pneumonia’. Br J Haematol. 2020;190(1):27–28.

30. Dzik S, Eliason K, Morris EB, Kaufman RM, North CM. COVID-19 and ABO blood groups. Transfusion. 2020;60(8):1883–1884.

31. Boudin L, Janvier F, Bylicki O, Dutasta F. ABO blood groups are not associated with risk of acquiring the SARS-CoV-2 infection in young adults. Haematologica. 2020.

32. Ward S, O’Sullivan J, O’Donnell JS. The relationship between ABO blood group, von Willebrand factor and primary hemostasis. Blood. 2020.

33. O’Donnell J, Boulton FE, Manning RA, Laffan MA. Genotype at the secretor blood group locus is a determinant of plasma von Willebrand factor level. Br J Haematol. 2002;116(2):350–356.

34. McKinnon TA, Chion AC, Millington AJ, Lane DA, Laffan MA. N-linked glycosylation of VWF modulates its interaction with ADAMTS13. Blood. 2008;111(6):3042–3049.

35. Levi M, Thachil J, Iba T, Levy JH. Coagulation abnormalities and thrombosis in patients with COVID-19. Lancet Haematol. 2020;7(6):e438–e440.

36. Murray GP, Post SR, Post GR. ABO blood group is a determinant of von Willebrand factor protein levels in human pulmonary endothelial cells. J Clin Pathol. 2020;73(6):347–349.

