## Supplementary material for "Blood group A Secretors are associated with a higher risk of COVID-19 cardiovascular disease complications": Mankelow et al 2020 Supplemental tables

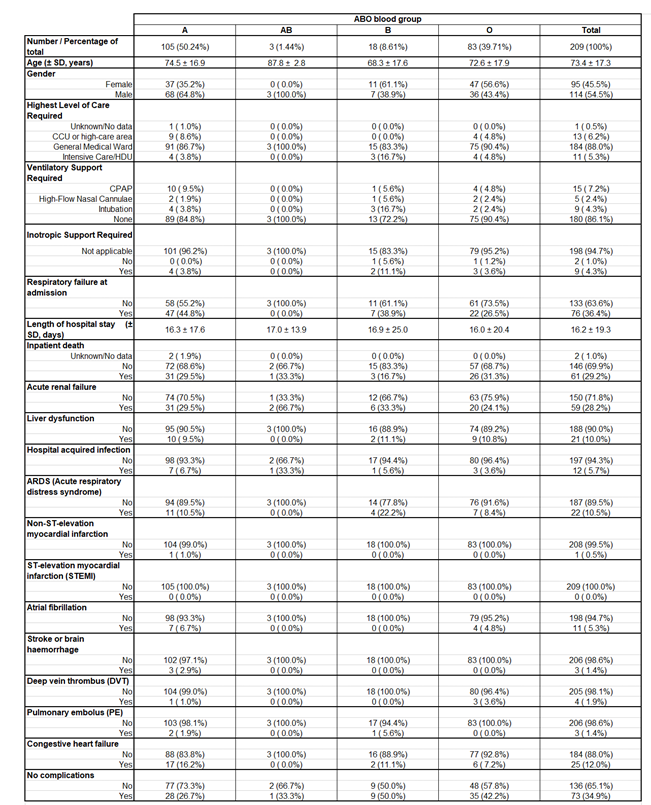

**Supplemental Table 1** (A more detailed version of table 1). A retrospective analysis of critically ill patients admitted to the intensive care unit at North Bristol NHS Trust (UK) with a positive PCR result for SARS-CoV-2 and for whom ABO blood group data was available.

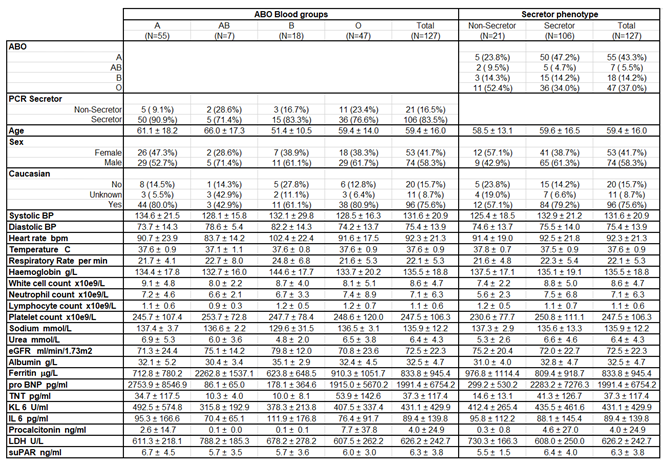

**Supplemental Table 2** - Retrospective analysis of patients admitted to North Bristol NHS Trust (UK) and enrolled onto the DISCOVER study for which ABO phenotype and full clinical measurements data was available. All clinical measurement were taken at the time of hospital admission

.

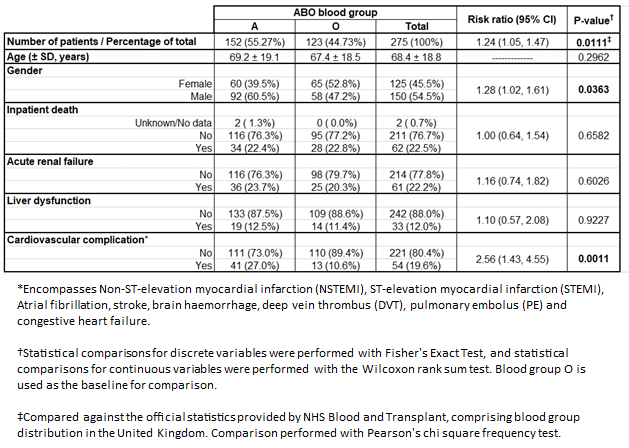

**Supplemental Table 3** – Statistical analysis of critically ill patients admitted to the intensive care in North Bristol NHS Trust (UK) with a positive PCR result for SARS-CoV-2 and that for whom ABO blood group data was available (Table 1) combined with data obtained through the DISCOVER study (Table 3).

-------------------------------------------------------------------------------------------------------------------------------------

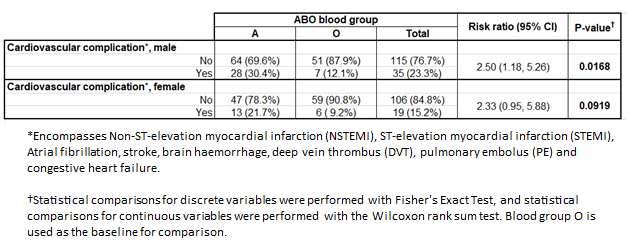

**Supplemental Table 4** – Statistical analysis of patients with cardiovascular complications, split into male and female, from supplemental table 3.
